# Prevalence of malaria among pregnant women attending ANC at public health institutions in Damote-Gale district, Southern Ethiopia

**DOI:** 10.64898/2026.09.21.26363605

**Authors:** Yeshake Kussa, Solomon Asnake

## Abstract

**Background:** In Ethiopia as other Sub-Sharan African countries malaria in pregnancy is a major public health threat particularly in the rural communities. Immunosuppression during pregnancy increases the risk of placental malaria; this, in turn, leads to anemia, low birth weight, preterm delivery, and stillbirth, causing severe complications that pose a life-threatening risk to both the mother and fetus. Hence, this study is aimed to assess the prevalence of malaria, and associated factors among pregnant women attending antenatal clinics in Damot Gale district, South Ethiopia.

**Method:** An institutional-based cross sectional study was conducted from March 1 to May 30, 2024. A multistage sampling technique and pre-tested semi-structured questionnaires were used for data collection. Using standard parasitological methods, blood samples collected from 389 participants were analyzed through microscopic examination of Giemsa-stained thick and thin blood films. Logistic regression was used to assess factors associated with malaria. Adjusted odds ratio with 95% confidence interval was calculated and P-value < 0.05 was considered statistically significant.

**Results:** In this study the prevalence of malaria among pregnant women was 10.5% (95% CI: 7.7–13.6%). Among the malaria-positive pregnant women (9%) were *P. falciparum* infected, followed by P. vivax (1.3) and mixed infections (0.2%). In the multivariate analysis, multigravdae (AOR = 0.358; 95% CI: 0.14–0.92), utilization of LLINs (AOR = 0.286; 95% CI: 0.119–0.687) and use of IRS (AOR = 0.068; 95% CI: 0.027–0.172) were significantly associated with malaria among pregnant women.

**Conclusion:** Malaria is still a public health problem among pregnant women in Damote-gale district. The present study indicated that adequate distribution and proper utilization of bed nets In addition seasonal proper spray of IRS might be helpful to prevent and control malaria.

## Back ground

Human malaria is caused by five primary protozoan species of the genus *Plasmodium*, transmitted via infected female *Anopheles* mosquitoes. The dominant species are ***Plasmodium falciparum*** (deadliest, most prevalent in sub-Saharan Africa) and ***Plasmodium vivax*** (widespread outside Africa), alongside ***Plasmodium ovale, Plasmodium malariae***, and ***Plasmodium knowlesi* [1]**. Despite millennia of efforts to combat malaria, the disease remains a formidable threat to global public health. The World Malaria Report 2025 reveals that the global burden continues to grow: an estimated 282 million cases and 610,000 deaths occurred in 2024 [2]. Sub-Saharan Africa bears the overwhelming burden, accounting for approximately 94% of all cases (265 million) and 95% of all deaths (579,000) in 2024. Five countries alone Nigeria, the Democratic Republic of the Congo, Uganda, Ethiopia, and Mozambique contributed nearly half of all global cases, underscoring the extreme geographic concentration of the disease [3].

In Ethiopia, Malaria peaks between April and May after the short rainy season (February to May) and between September and December after the long rainy season (June to September) [4]. In the country malaria is predominantly caused by two parasite species: ***Plasmodium falciparum*** (accounting for roughly 67% to 70% of cases) and ***Plasmodium vivax*** (contributing about 30% to 33%). Transmission occurs across roughly 75% of the country’s landmass, heavily impacting lowland and mid-altitude areas below 2,000 meters [5]. In 2023, 4.1 million malaria cases including 527 deaths were reported, of which *Plasmodium falciparum* accounted for 70% approximately of all reported cases. Additionally, there has been a shift in malaria stratification, with an increase in the number of areas classified as endemic in 2022 compared to 2020 [5].

Globally, an estimated 121.9 million pregnancies occurred in malaria transmission areas in 2020, with Sub-Saharan Africa accounting for approximately 46.1 million of these cases [6]. In 2021, in 38 moderate and high transmission countries in the WHO African Region, there were an estimated 40 million pregnancies, of which 13.3 million (32%) were exposed to malaria infection during pregnancy [7]. By WHO subregion, west Africa had the highest prevalence of exposure to malaria during pregnancy (40.7%), closely followed by central Africa (39.8%), while prevalence was 20% in east and southern Africa [8]. The expected pooled prevalence of malaria among pregnant women in Ethiopia was 12.72% [9]. Malaria during pregnancy might be asymptomatic due to a high level of developed immunity in mothers residing in high-transmission areas. However, it is still associated with an increased risk of maternal anemia, spontaneous abortion, stillbirth, prematurity, and low birth weight [10].

Damot Gale district is among the malaria areas in the South Ethiopia region. As a result, malaria prevention and control activities have been implemented among pregnant women. As to our review, there is limited evidence regarding malaria prevalence and its associated factors among pregnant women in the study area. In addition, Ethiopia has set goals for malaria elimination and conducting confirmatory testing for 100% of suspected malaria cases and treat as per the guide line is one of the main strategic objectives [11]. Therefore, this study aimed to assess the magnitude of malaria and identify its associated factors among pregnant women in Damot Gale district, Ethiopia. Thus, this study will provide valuable information to the district, zone, Region, the federal ministry of health, and different international partners on the magnitude of malaria and its associated factors among pregnant in the district that are important to target malaria control and prevention strategies and programs.

## Materials and methods

### Study area and period

A health facility based cross-sectional study was conducted at four health centers (Buge, Gacheno, Damot Mokonisa, Wandara gale) from January to May 2024 in Damot Gale district, which is located 350 km south of Addis Ababa Ethiopia. The area was located at 6°55’00” and 7°10’00” N Latitude and 37°45’0” and 38°0’0”E Longitude and at an altitude of 1500-2500m above sea level.. The average annual rainfall ranged from 1175mm to 1200mm, with an average temperature of 13.6 - 25.1 degrees Celsius. There are 32 villages with a total population of 143,705, consisting of 70,415 males and 73,290 females. According to Damot Gale district health office 2022/2023 season, about 4,972 pregnant women were estimated to give birth.

### Study design and study population

A health facility based cross-sectional study was conducted to determine malaria prevalence and associated factors among pregnant women at four health centers (Buge, Wandara, Mokonisa and Gacheno) in Southern Ethiopia. All pregnant women who attend antenatal clinic in the selected health centers at least for the last six months and willing to involve in the study during the data collection period were included in the study.

### Sample size determination and sampling technique

Using single population proportion formula with 50% prevalence, 95% confidence level, 5% margin of error and 10% non-response rate the total sample size was 422. However, the total number of symptomatic pregnant women in the study areas was 5170, which is less than 10.000. So, adjustment (correction) formula was used and 389 pregnant women have participated in the study. A multi-stage sampling technique was used to select pregnant women. Among 7 health centers, 4 were selected using simple random sampling technique. Then, the estimated sample size (389) was proportionally allocated to the selected 4 health centers based on the total number of 2738 pregnant women attending antenatal care in the selected 4 health center. Finally; using proportionate allocation, 192, 80, 52 and 65 study participants were involved from Buge, Wandara, Mokonisa and Gacheno health centers, respectively and enrolled by systematic sampling technique.

### Data collection procedure

Pretested structured questionnaires and checklists were used to collect socio-demographic, clinical, obstetric, and risk factor data from women who provided consent to participate in the study. Both data collectors and supervisors were trained for one day on all aspects of data collection procedures. The questionnaire and checklists were initially prepared in English and later translated to the local languages (Wolaitigna),) by native speakers of the languages. To ensure consistency, data collectors read and completed the local language (Wolaitigna) questionnaire for the participants, regardless of the participants’ literacy level. The data collectors’ supervisors and the principal investigator strictly checked the data on daily basis.

### Blood sample collection

For detection and identification of *Plasmodium* species, capillary blood was collected by trained and experienced medical laboratory technicians from each health center. Pregnant women’s finger was cleaned with 70% ethyl alcohol and the side of fingertip was pricked with a sterile lancet. The first drop of blood which contains tissue fluids was wiped away. One μl and 2 μl of blood were used for preparation of thin and thick blood films, respectively. Thick blood films were used for parasite detection and thin blood films were used for species identification. The prepared blood films were air dried and thin films were fixed with absolute methanol. The smears were then stained by 10% giemsa stain and examined under light microscope following standard operating procedures. A negative result was reported after checking at least 100 oil immersion fields. To ensure the quality of Giemsa stain, a quality giemsa stock solution prepared a head by expertise was used for preparation of 10% Giemsa (working solution) and prepared every 8 hours. Buffered water with PH-value of 7.2 was used for preparation of Giemsa stain working solution and filtered before use. At the end of data collection, all the slides were re-examined by experienced malaria microscopists in Sodo Hospital who have not been engaged during data collection. The re-examined slides was taken as final.

### Data management and analysis

Data was coded, entered, and cleaned using epi info version 7 and exported to the statistical package for social sciences (SPSS) version 26 statistical package for further analysis. Descriptive statistics findings were summarized using frequencies, percentages, tables, and graph. Binary logistic regression was performed to assess the predictors of malaria infection. In the bivariate analysis variables significant at *P* value of 0.2 were entered to multivariate logistic regression analysis model to control possible confounding effects and to determine factors associated with malaria. Adjusted Odds ratios (AORs) with 95% confidence interval were calculated and P-value < 0.05 was considered statistically significant.

### Ethical considerations

Ethical approval was obtained from College of Medicine and Health Sciences, Hawassa University, Institutional Review Board (Ref. No. IRB/171/16). Support letter was also obtained from Damot-Gale district health office and the four Health centers. Written informed consent was obtained from study participants after explaining the purpose of the study by data collectors. Study participants with positive results were treated according to the national malaria treatment guideline. All information obtained during the study was kept confidential.

## Results

### Socio-demographic characteristics of study participants

A total of 398 pregnant women participated in this study with a response rate of 100%. The mean age of pregnant women was 26.9 years (SD = ± 5.50) and the most predominant age group was 19-30 (70.9%). About 81.4% of the women resided in rural areas, 40.2% were illiterate, and 35.3% were engaged in day labor. Monthly income of the majority (82.9 %) was less than 1500 ETB. Majority (78.3%) of the pregnant women have family size less than or equal to 6. The majority (83.3%) of the study participants were prim and second gravidae whereas, about (76.6%) of them were in their first and second trimester of pregnancy.

**Table 1.** Socio demographic characteristics of pregnant women attending ANC at four health centers in Southern Ethiopia 2025 (n=389).

| **Variables** |  |  |
| --- | --- | --- |
| Age group | Frequency (n) | Percent (%) |
| 20-24 | 167 | 42.9 |
| 25-30 | 106 | 27.3 |
| 31-35 | 86 | 22.1 |
| 36-40 | 30 | 7.7 |
| Educational status |  |  |
| Illiterate | 157 | 40.4 |
| Primary | 125 | 32.1 |
| Secondary | 74 | 19 |
| College | 33 | 8.5 |
| Residence |  |  |
| Rural | 317 | 81.5 |
| Urban | 72 | 18.5 |
| Occupation |  |  |
| Daily laborer | 137 | 35.2 |
| house wife | 94 | 24.1 |
| Merchants | 85 | 21.9 |
| Civil servant | 73 | 18.8 |
| Monthly income |  |  |
| <700 | 168 | 43.1 |
| 700-1500 | 155 | 39.9 |
| >1500 | 66 | 17 |
| Family size |  |  |
| 1-3 | 155 | 39.8 |
| 4-6 | 111 | 28.5 |
| >7 | 123 | 31.7 |
| Gravidity |  |  |
| Multi-gravidae | 65 | 16.7 |
| Second-gravidae | 194 | 49.9 |
| Primi-gravidae | 130 | 33.4 |
| Gestation |  |  |
| 1st | 1st | 1st |
| 2nd | 126 | 32.4 |
| 3rd | 91 | 23.4 |

### Prevalence of malaria among pregnant women

The overall prevalence of malaria infection among pregnant women in the study area was 10.5% (95% CI: 7.7–13.6%). Among the malaria-positive cases, *P. falciparum* was the dominant species,accounting for 35 cases (9%), followed by P. vivax (1.3%) and mixed infections (0.2%). Malaria infection was highest among women aged 19–24 years, who accounted for 5.4% of the infected participants. In terms of monthly income, women earning ≤1,500 were more likely to be infected than those earning more than 1,500. Primigravidae had the highest prevalence of malaria infection (4.9%), compared with second-gravidae (3.9%) and multigravidae (1.8%). With regard to gestational age, malaria infection was more common among women in their first trimester (4.4%) than among those in their second (3.6%) and third trimester (2.3%) (Table 2).

**Table 2.**
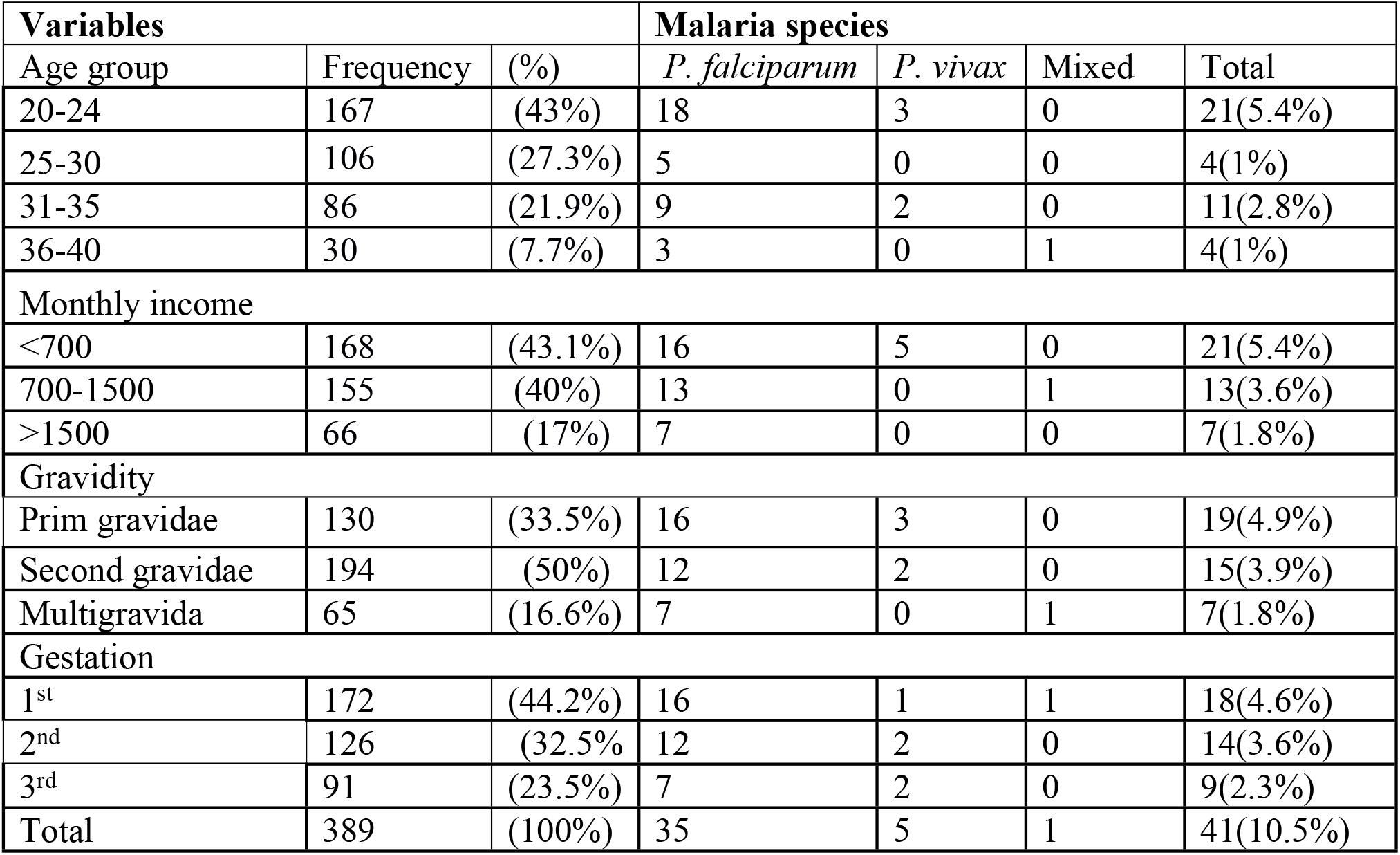
Prevalence of malaria among pregnant women attending ANC at four health centers in Southern Ethiopia 2025 (n=389)

### Knowledge level about malaria among pregnant women

About 355(91.3%) of the respondents had knowledge about malaria, health workers and community meetings being the main sources of information 365(93.8%). Knowledge of malaria transmission was also relatively high, as 315 (80.9%) of the pregnant women correctly identified mosquito bites as the mode of malaria transmission. Regarding malaria prevention, 75.3% (293) knew that LLINs protect against mosquito bites and contribute to malaria control. Most respondents 351 (90.2%) reported obtaining LLINs from public health facilities, while 320 (82.2%) reported using ITNs regularly. About 301(77.4%) of the respondents informed that their houses were indoor sprayed with chemicals in the last 12 months. Approximately 302 (77.6%) of respondents perceived IRS as advantageous, while 87 (22.4%) expressed concerns about its side effect.

**Table 3.** Knowledge level of malaria among pregnant women attending ANC at four health centers in Southern Ethiopia 2025 (n=389)

| **Variables** |  |  |
| --- | --- | --- |
| Way of Transmission | **Frequency (n)** | **Percent (%)** |
| Biting of mosquito | 315 | 80.9 |
| Airborne | 44 | 11.3 |
| Contaminated water | 30 | 9 |
| Protective measures |  |  |
| Use of LLIN | 293 | 75.3 |
| Cleaning environment | 70 | 18 |
| House cleaning | 26 | 6.7 |
| Sleep regularly under LLIN |  |  |
| Yes | 320 | 82.3 |
| No | 69 | 17.7 |
| Source of LLIN |  |  |
| Public health facility | 351 | 90.2 |
| Private health facility | 38 | 9.8 |
| Use of IRS in the last 12 months |  |  |
| Yes | 301 | 77.4 |
| No | 88 | 22.6 |
| Effect of IRS |  |  |
| Beneficial | 302 | 77.6 |
| Side effect | 87 | 22.4 |
| Information about malaria |  |  |
| Yes | 355 | 91.3 |
| No | 34 | 8.7 |
| Source of information about malaria |  |  |
| Community meetings | 202 | 51.9 |
| Health workers | 163 | 41.9 |
| Radio/Tv | 24 | 6.1 |

### Factors associated with malaria among pregnant women

Variables including age, educational status, occupation, gravidity, gestational age, use of protective measures, utilization of long-lasting insecticidal nets (LLINs), and indoor residual spraying (IRS) were significantly associated with malaria infection in the bivariate analysis. Variables with a *p*-value of less than 0.20 in the bivariate analysis were subsequently entered into the multivariable logistic regression model. In the multivariable analysis, gravidity, utilization of LLINs, and exposure to IRS remained significantly associated with malaria infection during pregnancy at a *p*-value of less than 0.05. Pregnant women who were multigravidae had 65% lower odds of malaria infection compared with primigravidae (AOR = 0.358; 95% CI: 0.14–0.92). Similarly, pregnant women who utilized LLINs had 71.4% lower odds of malaria infection compared with those who did not utilize LLINs (AOR = 0.286; 95% CI: 0.119–0.687). Furthermore, pregnant women living in houses that had been sprayed with IRS had substantially lower odds of malaria infection compared with those living in houses that had not been sprayed (AOR = 0.068; 95% CI: 0.027–0.172).

**Table 4.** Factors associated with malaria among pregnant women attending ANC at four health centers in Southern Ethiopia.

| Variables | Category | Malaria |  | COR (95% CI) | P.value | AOR (95% CI) | P.value |
| --- | --- | --- | --- | --- | --- | --- | --- |
|  |  | Negative | Positive |  |  |  |  |
| Age group (Years) | 20-24 | 146 | 21 | 1 |  | 1 |  |
|  | 25-30 | 101 | 5 | .580(.207,1.626) | 0.3 | .944(.281,3.170) | 0.926 |
|  | 31-35 | 75 | 11 | 2.217(.603,8.149) | 0.23 | 2.501(.580,10.794) | 0.219 |
|  | 36-40 | 26 | 4 | .482(.173,1.338) | 0.161 | .417(.125,1.397) | 0.156 |
| Education | Illiterate | 143 | 14 | .654(.141,3.027) | 0.172 | .852(.153,4.743) | 0.852 |
|  | Primary | 105 | 20 | .360(.079,1.631) | 0.185 | .453(.088,2.335) | 0.344 |
|  | Secondary | 69 | 5 | .890(.164,4.843) | 0.893 | .866(.144,5.209) | 0.875 |
|  | College | 31 | 2 | 1 |  | 1 |  |
| Gravidity | Primi gravidae | 111 | 19 | 1 |  | 1 |  |
|  | Secondi gravidae | 179 | 15 | .589(.228,1.521) | 0.274 | .449(.158,1.278) | 0.134 |
|  | Multigravida | 58 | 7 | .406(.173,.951) | 0.038 | .358(.140-.916) | .032* |
| Gestation | 1<sup>st</sup> | 154 | 18 | .377(.109,1.305) | 0.124 | .276(.068,1.124) | 0.072 |
|  | 2<sup>nd</sup> | 112 | 14 | .617(.169,2.253) |  | .341(.077,1.504) | 0.155 |
|  | 3<sup>rd</sup> | 82 | 9 | 1 |  | 1 |  |
| LLIN use | Yes | 309 | 11 | 1 |  | 1 |  |
|  | No | 40 | 29 | .099(.049,201) | 0 | .286(.119,.687) | .005* |
| IRS use | Yes | 293 | 8 | 1 |  | 1 |  |
|  | No | 56 | 32 | .054(.024,.123) | 0 | .068(.027,.172) | .000* |
| Protective measures | Use of LLIN | 231 | 18 | 1 |  | 1 |  |
|  | Env'tal cleaning | 69 | 11 | .354(.162,.773) | 0.009 | .713(.261,1.947) | 0.509 |
|  | House cleaning | 48 | 11 | .300(.131,.686) | 0.004 | .585(.190,1.803) | 0.351 |

## Discussion

In this study, the prevalence of malaria among pregnant women was found to be 10.5%, previous studies have reported varying prevalence rates and identified several factors that may influence malaria infection among pregnant women. The prevalence of malaria observed in this study was comparable with findings from South-West Nigeria, where a prevalence of 7.7% was reported [12], Southern Laos, where the prevalence was 8.3% [13] and a rural district surrounding Arba Minch Town, Ethiopia, where a prevalence of 9.1% was reported [14].

However, the prevalence found in this study was higher than that reported in Felege Hiwot Referral Hospital and Addis Zemen Health Center, Ethiopia (2.83%) [15], coastal Ghana (5%) [16] and India (5.4%) [17]. On the other hand, the prevalence was lower than research done at Pawe Hospital in Ethiopia, where the prevalence of malaria was (16.3%) [18] and Sudan (13.7%) [19]. The discrepancy of malaria prevalence between the current and previous studies might be due to the differences in the study periods. The present study was conducted from March to June which is the second-high malaria transmission season where high prevalence and epidemics occur in the country. However, prior studies were conducted in seasons where malaria transmission is considered low. In addition, this difference could be due to differences in sample size and socio-economic and demographic status of the study population included in the studies. The observed differences may be related to variations in geographical location, malaria transmission intensity, environmental conditions, and preventive practices across the study areas.

*Plasmodium falciparum* was the most predominant species with overall prevalence of 9 %, which is in line with the study done in Pawe Hospital, (9.67%) [18] and South Gonder and Baherdar northwestern Ethiopia (12.2%) [20]. But, this result is lower than the study conducted in Burkina Faso (26.9%) [21]. This variation might be due to the difference in inclusion criteria, study period and study site (type of health facility). The present study was conducted in health centers and pregnant women with severe malaria were excluded. However, Tahita and his colleagues’ study was conducted in hospital and included pregnant women with severe malaria which mostly occurs due to *P. falciparum*. In addition, the overall malaria burden is higher in Burkina Faso compared to Ethiopia as explained above.

The prevalence of *P. vivax* in the current study was 1.3% which is coherent with the study conducted in Pawe hospital (1.6%) [18]. However, this finding is lower than study conducted in North west Ethiopia (4.8%) and North Gonder, northwestern Ethiopia (3%) [22]. This variation might be due to variation in vector competence, rain fall, temperature and study site. The prevalence of mixed infections in the current study was (0.2%) which is lower than studies conducted in South Gonder, Baherdar (3.8%,) [20], North Gonder, (2.3%) [22] and Pawe hospital, northwestern Ethiopia (3.03%) [18].

Damot Gale District is located in an area where malaria transmission is relatively common, which may increase the likelihood of infection among pregnant women. In contrast, women living in areas with low malaria transmission have a lower risk of exposure and infection, which may result in lower prevalence rates. Therefore, differences in the epidemiological characteristics of the study areas may partly explain the variation in malaria prevalence between the present study and previous studies. Other factors, such as differences in study design, sample size, study period, diagnostic methods, and malaria prevention practices, may also contribute to the observed discrepancies.

Assessing pregnant women’s knowledge on malaria and associated factors is very helpful for policymakers and stakeholders in planning maternal and child health care services. In this study among women attending antenatal clinics 355(91.3%) had good knowledge on malaria. The finding of this study is comparable to studies done in Western Ethiopia 97.8% (436/446) [23] and study done in Cameroon (88%) [24]. However, the finding of this study is higher than a study done in Nigeria (83.9%) [25], Southern (73.4%) [26] and North west Ethiopia, (73.2%) [27]. Regarding malaria prevention, 293 (75.3%) knew that LLINs protect against mosquito bites and contribute to malaria control which is lower than finding of study conducted in North west Ethiopia 98.7% [27].

In the present study, socio-demographic, maternal, environmental, and personal factors were assessed. Gravidity, use of long-lasting insecticidal nets (LLINs), and use of indoor residual spraying (IRS) were found to be significantly associated with malaria infection.

Another important factor examined in this study was the relationship between gravidity and malaria prevalence. The findings of the present study showed that primigravidae pregnant women had higher odds of malaria infection compared with multigravidae women. This finding is consistent with studies conducted in southern Ethiopia [14] Nigeria [28], and the Congo [29], which also reported a higher risk of malaria among primigravidae women. Similar associations have also been reported in studies conducted in sub-Saharan Africa [30], Ghana [31], and Burkina Faso [21].

The higher risk of malaria among primigravidae women may be related to differences in immunity. Previous studies have shown that parasite density is generally higher among primigravidae and secundigravidae women than among multigravidae women. This may be because women in their first and second pregnancies have not yet developed sufficient pregnancy-specific immunity to effectively control malaria parasites, whereas multigravidae women may have developed greater immunity through repeated pregnancies and previous exposure to malaria.

However, the findings of the present study differ from those of Mohamud et al., who reported a higher malaria prevalence among multigravidae women (65.1%) compared with primigravidae women (34.9%) [32]. In addition, Yaro et al. (2021) [33] found no significant difference in malaria parasite prevalence among primigravidae, secundigravidae, and multigravidae women. Such differences between studies may be explained by variations in malaria transmission intensity, levels of exposure, immunity, and the characteristics of the study populations. In areas where all pregnant women, regardless of gravidity, are exposed to similarly high levels of malaria transmission, the likelihood of infection may be comparable across different gravidity groups.

The present study also demonstrated a significant association between the utilization of long-lasting insecticidal nets (LLINs) and malaria infection. Pregnant women who slept under LLINs had 71.4% lower odds of malaria infection compared with those who did not use LLINs. This finding is consistent with a study conducted in Benishangul-Gumuz, northwestern Ethiopia where pregnant women that slept under LLINs were 85% lower odds of malaria compered to those not used [34]. Similarly, another study conducted in North West Ethiopia found that pregnant women who used insecticide-treated bed nets were 87% less likely to develop malaria than those who did not use bed nets [35]. Studies conducted in in North Shoa Central Ethiopia [36] and Ghana have also reported that those who failed to use bed nets were 18.16 times more likely to be infected by plasmodium than bed net users [37]. associated with a substantially increased risk of Plasmodium infection. Other studies conducted in Ethiopia similarly found that bed net utilization was significantly associated with malaria infection, further supporting the importance of LLIN use as a malaria prevention strategy among pregnant women [38].

However, not all studies have reported a significant association between bed net utilization and malaria infection. Studies conducted in Burkina Faso [21] and Jawi District in northwestern Ethiopia [39] found no significant relationship between LLIN use and malaria prevalence. These differences may be explained by variations in the coverage, availability, and consistent utilization of LLINs across study settings. Higher LLIN coverage and regular use in some populations may reduce the observable differences in malaria infection between users and non-users. Differences in malaria transmission patterns, mosquito behavior, and other environmental and behavioral factors may also contribute to the variation in findings across studies.

The findings of this study showed that indoor residual spraying (IRS) was significantly associated with a reduction in malaria among pregnant women. Pregnant women living in houses that had been sprayed with IRS had 93% lower odds of malaria infection compared with those living in houses that had not been sprayed. This finding suggests that IRS plays an important role in protecting pregnant women from malaria infection. Our finding is consistent with previous studies conducted in different settings. For example, a study conducted in Pakistan reported efficient utilization of IRS reduced the incidence of P. falciparum by 98% and P. vivax by 90% [40], Uganda reported that the number of incident malaria episodes per person-week decreased by 83% following the implementation of IRS [41]. Similarly, a study conducted in Mozambique found that malaria prevalence was substantially reduced by 74% (95% CI: 72%–76%) after the implementation of IRS [42]. In Kenya, malaria prevalence decreased by 64.4% in intervention areas where IRS was implemented in valley regions after 12 months of follow-up, compared with non-intervention areas [43].

Similarly, evidence from Sub-Saharan Africa has demonstrated the effectiveness of reactive indoor residual spraying. A recent study reported a 68% reduction in malaria parasite prevalence following the implementation of reactive IRS [44]. These findings further support the important role of IRS as an effective malaria prevention strategy in malaria-endemic areas.

The findings of the present study are also consistent with evidence from Ethiopia. A study conducted in Western Ethiopia showed that IRS spraying of the pregnant woman’s house significantly reduced malaria prevalence. Pregnant women living in houses that had not been sprayed with IRS within the previous year had three times higher odds of developing malaria compared with those living in houses that had been sprayed. Other studies conducted in Ethiopia, including those by Gemechu et al. [45] and Tilahun et al.,[39] also reported a statistically significant association between the use of IRS and a reduction in malaria among pregnant women.

## Conclusion

This study showed that, malaria is still the major public health problem among pregnant women, though prevalence of malaria is decreasing in the country because of scale-up of intervention and prevention measures. *P. falciparum* was the predominant species causing malaria infection in the study area. Primigravidae, not sleeping under LLINs and living in houses not sprayed were factors that increased the prevalence of malaria infection in pregnant women. Therefore, prevention and control of malaria strategies should be strengthened and community-level screening of pregnant women and community level focused antenatal services for pregnant women might be important to avoid malaria infection during pregnancy.

## Acknowledgements

The authors are grateful to the study participants for voluntary involvement in the study and providing the required information. The authors would like to thank Damote-Gale district health offices and respective health center leaders and staffs for their support throughout the investigation providing the required information. We acknowledge Hawassa University and for providing supply for data collection only. We would also like to extend our heartiest appreciation to data collectors too.

## Declarations

We declared that this article is our original work, and this manuscript is not submitted to other journals elsewhere.

## Data availability

The data that support the findings of this study are available from the corresponding author upon a reasonable request.

## Authors’ contributions

YK: drafted the proposal and SA reviewed it. YK collected the required data and analyzed it. SA prepared the manuscript and both authors read and approved the final manuscript

